# Vaccine attitudes and perceptions in a post-COVID Era: what lessons can be learned in Tunisia?

**DOI:** 10.64898/2026.10.02.26364577

**Authors:** Nadia Ben Mansour, Beya Farah Benzina, Saoussen Fray, Emna Hariz, Ahlem Gzara, Habiba Ben Romdhane, Judith E. Mueller, Slimane Ben Miled

**Affiliations:** MedicalFaculty of Tunis, Tunisia; Institut Pasteur de Tunis, Tunis, Tunisia; Faculté de sciences humaines et sociales de Tunis, Tunis, Tunisia; Ministry of Health, Tunis, Tunisia; Ecole des Hautes Etudes en Santé Publique, Rennes, France

**Keywords:** Vaccine hesitancy, Human papillomavirus, COVID;Low and Middle income countries, Muslim countries

## Abstract

**Introduction:** Although the COVID-19 pandemic clearly showed the potential consequences of a world without vaccinations, vaccine hesitancy is likely to be more pronounced in the post-COVID-19 era. The introduction of human papillomavirus (HPV) vaccines in this context is likely to raise particular concerns. In Tunisia, decision-makers have prioritized the introduction of HPV vaccine into the immunization schedule. This study assessed individual barriers and facilitators to HPV vaccine acceptability in Tunisia, with the aim of guiding an effective communication strategy for HPV vaccination campaigns in Tunisia and neighboring countries in a post-pandemic context.

**Methods:** Using a qualitative design, we conducted 14 focus groups (FG) with healthcare professionals (HCP) (N=32) and community representatives (CR) (N=89) in five regions of the country. The discussion followed a semi-structured guide designed on the basis of vaccine hesitancy determinants matrix. Transcripts were deductively analyzed according to the “social processes” and “thinking and feeling” of the behavioural and social drivers (BeSD) Framework. Inductive approach was used to analyze participants’ suggestions for promoting HPV vaccination.

**Results:** The FG analysis demonstrated that the historically strong trust in vaccination in Tunisia has weakened after the COVID-19 pandemic. Mistrust of health system and experts, the pandemic management and vaccine mandates measures had negative influence on the vaccine attitudes in a post pandemic Tunisian context. Persistent concerns about COVID-19 vaccines safety were frequently intertwined with widespread distrust of pharmaceutical companies and their profit motives, perceived as imposing inequitable access and lower-quality vaccines on countries in the Global South. These concerns fueled confusion about vaccine’s side effects. The weakening of immunity was recurring, as well as memory issues, impacts on fertility, and sudden deaths. Regarding information, our research revealed that although digital platforms were perceived as widely used, a significant part of the population interviewed considered public TV channel, as the most reliable source of health information.

**Conclusions:** Emphasizing how the pandemic has shaped individual and collective perceptions and attitudes toward vaccination, our findings underline the importance of leveraging lessons learned from the pandemic to develop a context tailored and coherent communication strategy for successful HPV vaccine implementation.

## Introduction

Health systems were globally reshaped by the COVID-19 pandemic [1]. Despite the impressive and rapid scientific and technological innovationof vaccines manufacturing that had significantly changed the course of the pandemic [2], immunization coverage has still not returned to 2019 levels in many countries [3,4], creating immunity gaps worldwide. Several factors have been incriminated, including vaccine hesitancy [5]. Vaccination hesitancy defined as “as a state of indecision and uncertainty that precedes a decision to become (or not become) vaccinated” [6] becomesmore than ever, a significant threat to public health [7].Indeed, individuals who were skeptical about COVID-19 vaccines may generalize their distrust to other vaccines [8].

In Tunisia, an Arab country within theEMR, vaccination plays a key role in the health system. The National Immunization Program (PNV) covers most of the vaccines recommended worldwide. However, contextual variability and budgetary constraints account for the absence of certain vaccines, such as those for rotavirus, HPV, and pneumococcus [9].The introduction of the HPV vaccine is plannedfor 2025, targeting girls in their sixth year of primary school, as announcedby the Ministry of Health [10]. Alongside cultural factors, an emerging vaccine hesitancy has been observed in Tunisia following the COVID-19 pandemic, mirroring trends seen in many other countries [11,12].

Achieving the WHO target of 90% HPV vaccine coverage [13], requires addressing vaccine hesitancy, which are inherently complex [14].As reported in a large number of studies from several continents, individual attitudes are among major obstacles to the vaccine uptake [7,15].Notablefeatureof these attitudes includes the lack of trust in vaccine safety and efficacy, with concerns about potential side effects or long-term consequences of vaccination [16,17]. Such fears are frequently exacerbated by misinformation circulating on social media and other platforms, further shaping perceptions and decision-making [18].

Another key factor reported in the literatureis the perception of risk. Some individuals underestimate the seriousness of vaccine-preventable diseases or overestimate their own health, fostering a sense of complacency [16,18]. This distorted risk perception may lead to viewing vaccination as unnecessary on a personal level, even when its broader public health value is acknowledged. Trust in healthcare providers, scientific institutions, and government authorities also plays a pivotal role in shaping these attitudes [18].When trust is low, people are more likely to be skeptical of vaccine recommendations and may seek alternative sources of information, which can further reinforce hesitancy [19].Personal experiences and cultural beliefs can significantly influence individual attitudes as well. For example, previous negative experiences with healthcare systems or vaccines may contribute to hesitancy [16,20], underscoring the importance of considering past vaccine experiences in tailoring communication efforts for future vaccination.

In this particular context, we aimed to analyze attitudes and perceptions of HPV major stakeholders in Tunisia (community and healthcare professionals) regarding vaccination in the post COVID era, in order to shed light on links between COVID-19 vaccine hesitancy and HPV vaccine acceptance.

## Method

### Study design

The institutional review board of Pasteur Institute of Tunis approved the study protocol (2023/14/I/V2; 13 December 2024). Using a concurrent mixed-methods approach with a dominant integrated qualitative design [21,22], we conducted 14 focus groups (FG) discussions sessions combined with brief surveys over the period between September 2023 and February 2024. The qualitative study aims to explorethe attitudes and perceptions with regard to vaccination, and the quantitative parthas prioritized a set of 7 communication attributes related to HPV vaccine,andidentified through a literature review [23,24].The prioritized attributes will be further tested using a discrete choice experiment (DCE) study [25,28]. Communication attributes rating and DCE results will be reported elsewhere.

Purposive cluster multistage sampling was used to understand both variations and similarities [22],of vaccine acceptance among demand’s HPV vaccine stakeholders, namely healthcareprofessionals (HCP) and precisely parents and school staff, called thereaftercommunity representatives (CR) [29,30].

### Recruitment

For the first cluster, five groups of HCP were identified as the most involved in HPV vaccine counselling: Midwives, Gynecologists, pharmacists,pediatricians and general practitioners [31,32]. Key informants (n=32) from both public and private sectors were identifiedthrough relevant academic societies and professional associations networks 80.

For the CR cluster, maximum variation sampling according to geographic area was used to account for disparities in social health determinants [33].Otherwise, the participants recruitmentwas undertaken by National Office of the Family and Population (ONFP) headquarters’and regional staff.Individuals who are in daily contact with children and teenagers and are involved in their health-related decision-making, such as parents, teachers, or any other child related profession (childcare educators, kid’s entertainers…)

### Data collection

A semi-structured discussion guide was designed based on vaccine hesitancy determinants matrix [34,35], that was successfully applied to analyze COVID19 vaccine hesitancy [36]. FG discussions were structured in three parts: 1) contextual influences and confidence in health system and institutional performance before and after the pandemic, 2) personal and group experience with vaccination3) Knowledge and perceptions specific to genital cancer and HPV, and information needs (FG guide is attached as appendix). This paper will report the results related to the two first parts.

The discussion was moderated by a socio-anthropologist [SF] and a medical doctor [BFZ]. Each session began with an introduction to the purpose of the FG and an explanation of the confidentiality of the discussions.Written informed consent was obtained.

### HCR’s FG discussion sessions

Initially, four focus groups were organized with HCP, each representing a specific specialty, and one group combining pediatricians and general practitioners. The sessions were held in the premises of each related scientific society or professional association in the capital Tunis.

### CR’s FG discussion sessions

Two gender-stratified FGs with CR were held per region, in the premises of the ONFPlocated in 3 coastal and 2 inland areas, spread over the North, Center and the South of the country (see table 1). This choice reflects the socio-economic and cultural disparities along North/South and Costal/inland axes [37].

Table 1. Geographic distribution and dates of Focus groups with CRs.

Each session started with an introduction to the purpose of the FG and an explanation of the confidentiality of the discussions. After obtaining the agreement of the participants, the discussionwas audio recorded. Participants had not received any HPV vaccine-related pre-sensitization.

The duration of each FG was an average of 120 minutes. The discussion was moderated by the same duo as for the FGs with professionals, according to the same discussion guide.

### Data analysis

FG discussions were transcribed by SF and thematically analyzed [38,39]by NBM and BFZ research team who independently reviewed transcripts and agreed on a consensus recurring categories and variants according to predefined categories from the Behavioral and Social Drivers of Vaccination Framework (BeSD)[40,41].The analysis focused specifically on the “social processes” and “thinking and feeling” (see figure below).

All pertinent transcript data was manually inputted, to the coding frame by topic and categories using an excel codebook. Patterns of agreement and similarities across groups were then examined during in person meetings. The analysis was stratified according to specialty for HCP’s FG, and sociodemographic characteristic for CR’s FG (gender, region, occupation). The quotes cited in this publication, were translated to English by NBM.

Concerning the communication attributes ranking analyses, the statistical analysis was conducted using SPSS software, version 21 (Version 21.0. Armonk, NY: IBM Corp). The top-ranked attribute was identified by computing percentages of participants’ who ranked first the attribute.

## Results

### 1. Socio-Demographic Characteristics

A total of 32 HCP participated in 4 FG, one third of them (11) being midwifes (figure 2)

**Figure 1.**
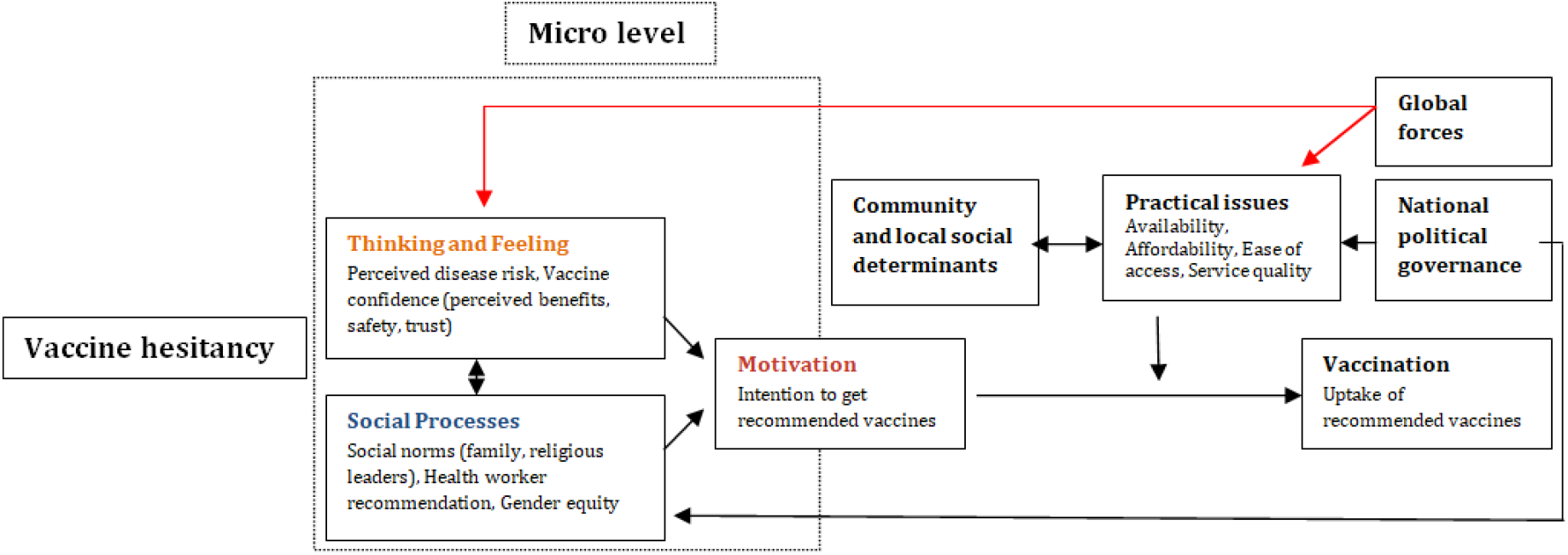
Framework of Vaccine Hesitancy Determinants and Vaccination Uptake (adapted from WHO SAGE BeSD Framework).

**Figure 2.**
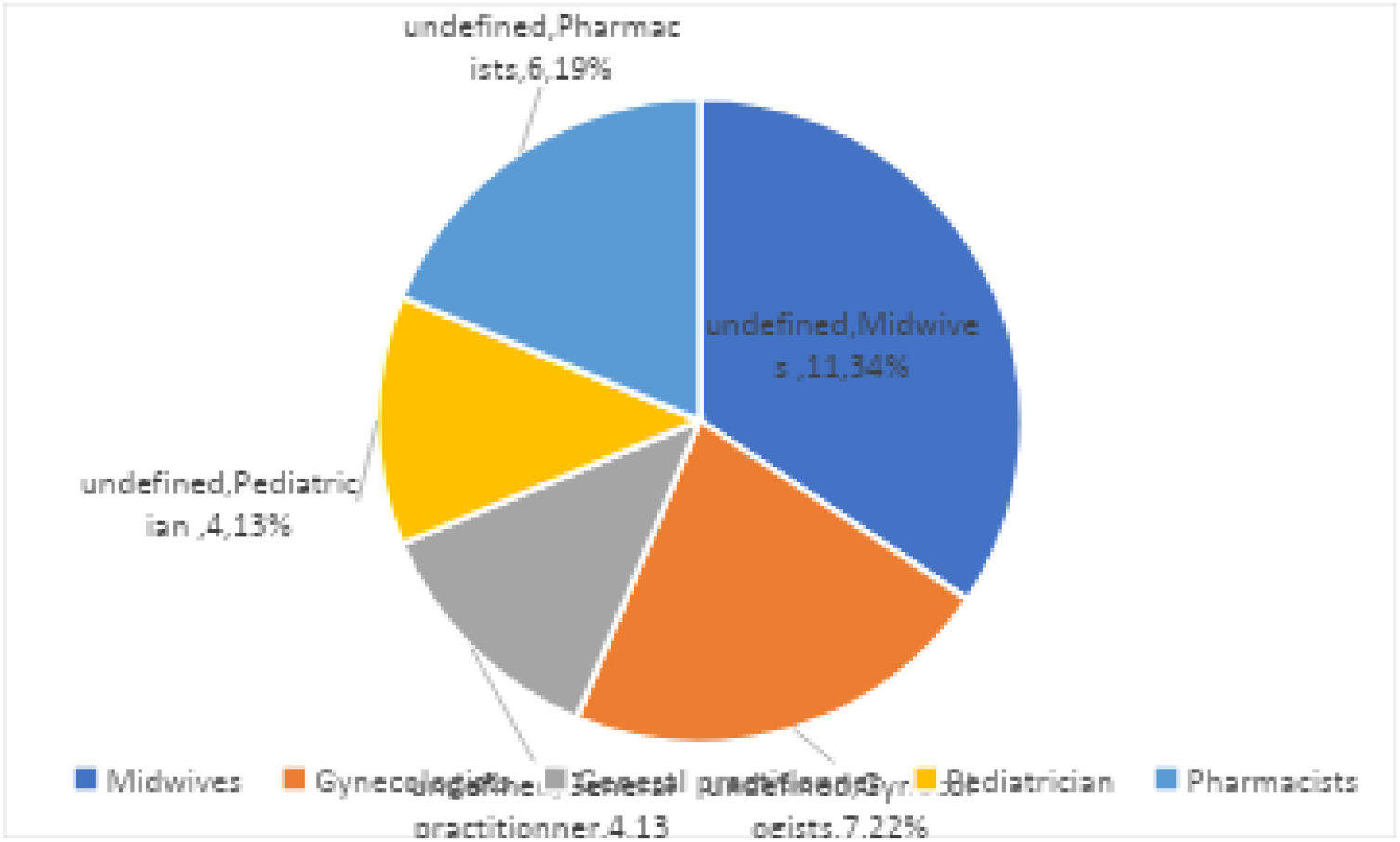
Distribution of HCPs participantsby category.

Ninety-four community representatives participated in ten focus group discussions, with distinct sessions for men and women in each region. The participants were mainly aged 30-50 years (Table 2).

**Table 2.** socio-economic characteristics of Community Focus Group participants.

| Focus Group | Sousse | Jendouba | Tuniss | Gafsa | Médenine | Total |
| --- | --- | --- | --- | --- | --- | --- |
| **Gender** |  |  |  |  |  |  |
| Men | 9 | 9 | 7 | 9 | 9 | 43 |
| Women | 11 | 9 | 12 | 9 | 10 | 51 |
| **Age** |  |  |  |  |  |  |
| <30 | 2 | 2 | 4 | 2 | 0 | 10 |
| 30-50 | 14 | 11 | 11 | 13 | 15 | 64 |
| >50 | 4 | 5 | 4 | 3 | 4 | 20 |
| **Role** |  |  |  |  |  |  |
| Parent | 9 | 13 | 13 | 8 | 10 | 53 |
| Teacher | 9 | 3 | 3 | 8 | 8 | 31 |
| Child focused profession | 2 | 2 | 3 | 2 | 1 | 10 |

### 1. Insights from Discussions

Seven major themes had emerged from our analysis of the FG data: 1) Male authority and fatalism had negative influence on the vaccine uptake during the pandemic, 2) historically strong trust in vaccination in Tunisia is being increasingly questioned after the political uprisal and the pandemic3) mistrust of health system and experts following the pandemic management and scientific uncertainty; 4) suspicions about the role of the pharmaceutical industry, 5) concerns about the safety and long-term effects of the vaccines 6) Polarizing vaccine mandates measure debate 7) Trusting TV as a source of medical information.

#### Theme 1: Gender roles and fatalism rise to the surface

Tunisian culture, significantly influenced by traditions and social norms, has a considerable influence on vaccination practices, particularly with regard to gender roles. Testimonies from NW and SE, stated thatit is “naturally” women who are tasked with taking children for vaccinations, a responsibility seen as “*more closely linked to the mother*” because of entrenched social belief that parenting abilities are tied to one’s gender (see supplementary table 1, Q1 and Q3).

Nevertheless, in the capital, one participant expressed a differing opinion reflecting a willingness to take his children for vaccinations, indicating a more progressive cultural attitude in the capital relative to the more conservative areas in the south and the interior of the country (see supplementary table 1, Q2).

According to participants prevailing views, women have the responsibility for accompanying children, while men claim control over decisions regarding their wives and children, notably in the context of COVID-19 vaccination, *[prevented them from getting vaccinated against COVID]*, according to a father from SE (see supplementary table 1, Q4)

Fatalism reinforcedby unwavering belief in fate “*God has already decided my fate and my death*” and lack of trust in the state, had also a significant role in shaping individuals’ perceptions and opinions regarding vaccination acceptability. Testimonies from the southern regions illustrate these perspectives (see supplementary table 1, Q5 and Q6).

#### Theme 2: Historical Influences: vaccination as social norm

We noticed a high level of trust in the national vaccination program, perceived as a “success” by various participants across the country, including both professionals and a mother from NW (see supplementary table 1, Q7 and Q8).

This trust is rooted in the perception of the State, embodied by President Habib Bourguiba, as a provider of essential services and a builder of the future for next generations, as it was mentioned by a father from the capital Tunis (see supplementary table 1, Q9).

Another father from the SE highlighted how much the vaccination practices are deeply embedded in the collective memory of Tunisians, evolving into a widely accepted social norm that perpetuated the transfer of these practices across generations (see supplementary table 1, Q10).

On this point,a participant from CE highlighted that vaccination in Tunisia is more of a “*deep-rooted habit”* than a sign of trust (see supplementary table 1, Q11).

In a similar spirit, a teacher from SE stressed that significant events in Tunisian history, such as the Revolution of 2010 and the COVID-19 pandemic, have challenged this perception and raised doubts about the State’s role as a welfare State but in the context of *“enlightened despotism”*, claiming a certain level of self-determination in line with the Revolution values (see supplementary table 1, Q12).

#### Theme 3: Mistrust of the Health System and experts

The health crisis significantly weakened the public trust in the healthcare system, as illustrated by the testimony of a teacher from the capital, who expressed her diminished confidence in the Ministry of Health following its response to the pandemic (see supplementary table 1, Q13).

Another mother from SE criticized the state’s response with regards to curative and preventive care of COVID 19, the health system was thought to be directly responsible for the deaths of thousands of people (see supplementary table 1, Q14).

In discussions about vaccination, a midwife from the capital made a clear distinction between the pre- and post-COVID periods, emphasizing how mistrust had affected people’s willingness to receive vaccines (see supplementary table 1, Q15).

This environment of mistrust was further exacerbated in the inland regions (SW and NW), where teachers reported parental refusal of routine childhood vaccination (see supplementary table 1, Q16 and Q17).

This statement was confirmed by a mother from SW who stated that she chose not to vaccinate her children. She explained that the informational document about vaccines had only fueled her doubts, prompting her to decline vaccination (see supplementary table 1, Q18).

Although previous statements highlited childhood vaccination refusal, a pharmacist from the capital offered a contrasting perspective, indicating that vaccination rates were predominantly impacted among adults than among children (see supplementary table 1, Q19 and Q20).

#### Theme 4: Blaming the pharmaceutical Industry

The COVID-19 pandemic intensified debates about the role of the pharmaceutical industry, particularly regarding the vaccines’ security, as it was highlighted by many participants from different Regions. According to teacher from the CE, this industry, faced allegationssuggesting the virus itself being created in pharmaceutical labs (see supplementary table 1, Q21).

Furthermore, Western laboratories were perceived as imposing inequitable or lower-quality solutions on countries in the Global South. Two mothers from the south made reference to abusive and unethical experimental practices *“laboratory rats”* (see supplementary table 1, Q22 and Q23).

In contrast, there was greater trust expressed regarding potential domestic products. As the mother from SW expressed (see supplementary table 1, Q24).

During a discussion with healthcare professionals, a pharmacist shared the previously expressed apprehensions, particularly regarding the financial intentions of pharmaceutical industries (see supplementary table 1, Q25).

#### Theme 5: Concerns about the safety and long-term effects of COVID-19 Vaccines

Fear of contracting COVID-19 was frequently cited as the main reason for vaccination, a perspective echoed by both community and healthcare participants (see supplementary table 1, Q26 and Q27), yet concerns about vaccine safety emerged as an even more prominent factor mainly attributed to the rush in its commercialization, as indicated by communication and education professionals(see supplementary table 1, Q28 and Q29)

Furthermore, the lack of information and uncertainty about the side effects of the COVID-19 vaccines were evident during the discussions. A teacher from NW expressed criticism towards healthcare professionals for emphasizing the benefits of vaccination while overlooking its associated risks (see supplementary table 1, Q30).

The lack of long-term surveillance also raised some doubts among health professionals, leading to conflicting statements. Examples of contradictions *“conflicting opinions”* were emphasized during discussions, deepening uncertainty about the safety of the vaccines, as a mother from CE noted (see supplementary table 1, Q31).

The geopolitical context was also pointed by a teacher from NW as a contributing factor to the prevailing uncertainty among experts. According to teacher from NW, skeptical perceptions of Chinese and Russian vaccines, were shaped by ongoing geopolitical conflicts with the United States (see supplementary table 1, Q32).

Overall, safety concern fuled by perceived inconsistencies in scientific evidence contributed to participants’ confusion between COVID-19 symptoms and potential vaccine side effects. Reported weakening of immunity was recurring, as well as memory issues, impacts on fertility, and sudden deaths (see supplementary table 1, Q33, Q34, Q35 and Q36).

#### Theme 6: TV is still the most reliable information’s source

Understanding participants’ concerns and uncertainties also requires examining the health information sources they relied upon. Our research revealed that although digital platforms are widely used, particularly among young people, a significant part of the participants interviewed still perceives the public TV channel “El Wataniya 1”, as the most reliable source of health information (see supplementary table 1, Q37 and Q38).

For instance, a midwife stated that TV is also the most accessible chanel for health information, especially in areas where internet access is limited (see supplementary table 1, Q39), in contrast to digital social networks, which a teacher from SE questioned for its credibility (see supplementary table 1, Q40)

Nevertheless, the trust in national channel has been put into perspective by a father from SE, highlighting that the credibility of information depends more on the presenter than the media itself (see supplementary table 1, Q41)

#### Theme 7: Vaccination mandates is a polarizing measure

Dissatisfaction with the mandatory vaccination policy was salient during the discussions, A father from NW highlighted the lack of clear information regarding its benefits and risks, which led to the perception of vaccination as an imposition rather than an informed decision (see supplementary table 1, Q42).

In reaction to these statements, many participants conveyed a deep sense of responsibility towards the healthcare system, which they believed would struggle to cope with a massive influx of sick people (see supplementary table 1, Q43 and Q44).

A mother from SE strongly suggested to make the introduction of new vaccines mandatory, referring to the current skepticism of parents in relation to vaccines in post COVID era (see supplementary table 1, Q45).

## Discussion

We used a qualitative design to analyze attitudes and perceptions of HPV major stakeholders (community and healthcare professionals) regarding vaccination in the post-COVID era, in order to help tailoring communication strategy around HPV vaccination. Making use of information from several regions and sources, our study had allowed a novel and thorough understanding of the pandemic impact on perceptions of Tunisian adults and HCPs regarding vaccination. Given the very low coverage of the HPV vaccine following its introduction in Tunisia in April 2025, our findings are particularly relevant for national health authorities. Furthermore, these results may offer insights applicable to other Arab or Muslim countries that share similar historical and socio-cultural contexts, helping to understand potential drivers of HPV vaccine hesitancy beyond Tunisia.

Our findings pointed out seven themes: 1) Male authority and fatalism had negativeinfluence on the vaccine uptake during the pandemic 2) Historically strong trust invaccination in Tunisia is being increasingly questioned after the political uprising (well knownJasmine Revolution) and the COVID-19 pandemic, 3) Mistrust of health system and expertsfollowing the pandemic management and scientific uncertainty; 4) Suspicions about the roleof big pharmaceutical industry 5) Concerns about the safety and long-term effects of COVID-19 vaccines 6) Polarizing vaccine mandates measure debate 7) Trusting TV as a source ofmedical information. The Three first themes are closely linked to the Social Processes’ domain of BeSD Framework, and the four others relates to “Thinking and Feeling”issues. These results highlight how the pandemic has affected individual and collective experiences and attitudes toward vaccination. We must also acknowledge, that despite its importance,practical facilitators such convenience and vaccine access did not emerge as a concern in our study.

Since COVID -19 pandemic, an abundant body of literature emerged on the subject of theattitudes and perspectives regarding COVID-19 vaccine.Recent studies [42-45], among which systematic reviews [14,43] confirmed that the lowest acceptance among adults was found in the Middle East countries and Muslim communities. This fact may impede the introduction of a new vaccine among these populations. A review article on the determinants of vaccine hesitancy in Muslim countries [46] had identified religious and cultural beliefs, as well as misinformation and conspiracy among its main drivers.

Our results mirrors in many aspects those of a Malaysian study, with regard to the distrustin healthcare system and pharmaceutical industries, religious fatalism, security concerns and misinformation with regard the COVID vaccines [42]. Widespread conspiracy theories, withvaccines being perceived as Western thread, had also compounded measles vaccinehesitancy in Sudan [47]. Historical injustice was also a recurring theme when studyingvaccine hesitancy among African American population (48,49), in Sierra Leon [50]wherecolonial narratives influenced Ebola vaccine hesitancy and in Nigeria,where the polio vaccine was boycotted due to previous unethical pharmaceutical trials [51].

Historical dimension was strongly present in our data, and fueled both positive and negativeperceptions. A historical trust in the national vaccination program was based on the idea thatthe State, perceived as the symbol of the independence,was committed to safeguarding thepopulation’s well-being. Nevertheless, our study highlights that key historical events, such as the 2010 Revolution and the COVID-19 crisis, have eroded this perception and called intoquestion the role of the State as a “welfare state”. The criticism of these policies is marked inthe southern regions (Medenine) and the interior regions (Gafsa and Jendouba), whereparental refusal of vaccination has been reported. Such a result could be explained bycollective frustration with regards to public policies that have failed to address persistentdisparities between coastal areas (better-off) and the interior (poorer regions like the Centerand south West) [52,53]. An ethical debate on the vaccine mandate were also risen duringthe discussions, which was experienced as a wound by some, while others (withcertain utilitarian philosophy) perceive it as a solution to counteract vaccine hesitancy.The decline in trust in Tunisia’s vaccination programs in post-COVID-19 is not a Tunisianparadigm. Globally, COVID-19 highlighted systemic fragilities, as demonstrated by recentstudies from high income countries which discussed how trust in health systems can weakenunder crisis conditions [40,54]. A study conducted in the United States, betweenApril 2020 and January 2024, found that trust in HCP and healthcare systems decreased throughout the pandemic across all sociodemographic groups and that it was associated with decreased likelihood of vaccination against COVID as well as influenza [55]. However, this distrust is particularly amplified in Tunisia by political and historical factors, above all the Western past colonization. The perception of being an “experimental animal in a laboratory” reflects a persistent sentiment among part of the Tunisian population of ongoing exploitation by developed countries.Unexpectedly, this conspiratorial discourse was shared by some HCP, notably pharmacists. Similar findings were reported among pharmacists and other HCP elsewhere, highlighting the lack of information of HCP about the vaccine’s security [56-58].Misinformation about adult’s vaccine’s security is a continuous feature in Tunisia, and was reported in a 2019 study about the influenza vaccine perceptions. It was even reported that some received advice against its uptake from HCP [5].The COVID-19 pandemic intensified debates about the role of the pharmaceutical industry,particularly regarding the production and distribution of vaccines. According to a 2022economic analysis [60], the global intellectual property (IP) framework promotes industrialmonopolies and exacerbates distribution inequity. International initiatives such COVAX failedto address the South-North inequity since it relied on public-private partnership. Indeed,through bilateral agreements, wealthy nations monopolized vaccination supplies, denyinglow- and middle-income countries (LMICs) access to necessary doses [60].

This industry, already viewed with suspicion by many, faced allegations suggestingthe virusitself being created in pharmaceutical laboratories [61], as reflected in our findings.Conspiracy theories and rumors are intensified through digital platforms, exacerbatingclimate of distrust and vaccine’s hesitancy [62,63]. Many participants in our study wereaware that some online platforms frequently magnify health information and disseminatemisinformation.A recent analysis [64], suggests that vaccine hesitancy is less aboutmisinformation and more rooted in distrust of authorities, institutions, and experts. Ratherthan stemming from a lack of knowledge or concerns about the vaccine itself, hesitancy isoften driven by deeper issues of institutional mistrust. Authors emphasized the relevance ofunderstanding collective offline experiences, as well as broader historical and politicalcontexts, to better understand how individuals interact with digital (mis)information and howit influences their attitudes and decision making on vaccinations.

### Limitations

This study has mainly two major limitations. First, the FGs included working age groupadultsso attitudes of younger age groups may have different experiences and sources ofinformation. Thus, the future quantitative study will focus on young people to understandtheir preferences towardvaccines.

Second, due to qualitative sampling procedures and social desirability bias interpreting and generalizing the results should be done with caution, particularly for segments of professionals and to individuals with higher socioeconomic status. Future longitudinal and cross-sectional survey data could overcome these gaps.

### From COVID-19 to HPV: Implications for Future Vaccine Acceptance

As reported elsewhere [65], mistrust in experts, healthcare system, state and pharmaceutical laboratory was highlighted in our data as an underlying cause of vaccine’s negative attitudes.

Addressing the complex issue of mistrust, require multiple strategies. In addition to usingsocial listening to address specific concerns [66], lessons from African countries [67]demonstrated the relevance of community engagement and collaboration with trustedindividuals or organizations to promote vaccine messages.

Effective communication emerged also as a pivotal factor in improving HPV vaccineacceptance across Kenya, India, and Nigeria [68] in post COVID era. Healthcare workers, as trusted sources, were trained not only in disseminating vaccine knowledge but also adapting an empathetic, culturally sensitive communication strategies to address concerns and counter misinformation. Multisectoral collaboration, including education, media, andcommunity leaders further strengthened outreach and fostered trust-building. Framing thevaccine as a meanfor cancer prevention, demonstratinglong term efficacy and security,proved particularly persuasive.

Secondly, media exposure also appeared to be a key factor influencing perceptions and attitudes toward vaccination. In this study, television was identified as the primary source of information among participants. However, social media platforms must also be carefully considered in the design of communication strategies, given their documented role in the amplification of anti-vaccination narratives across multiple settings, including Tunisia [69,70].

Above all, the recent global resurgence of vaccine hesitancy is certainly linked to the COVID-19 pandemic, but the rapid expansion of social media also played a major role in this phenomenon, as explained by Chen Yin [69]. Therefore, a strong communication strategy to address HPV vaccine hesitancy must effectively leverage both traditional media and social media platforms.

Another important aspect to consider for communication strategies is the heterogeneity of the audience. Various degrees of opposition to vaccination exist, ranging from vaccine hesitancy to outright refusal [71]. This polarization was also observed in our findings through participants’ perspectives. Our results further highlight a geographical dimension to this polarization. Participants from southern and interior regions appeared more critical and expressed lower levels of trust in governmental decisions compared to those from coastal areas. This regional disparity emphasizes the need for context-specific and targeted communication strategies.

Evidence from previous implementation experiences underscore that successful HPV vaccination programs depend as much on strategic communication as on operational logistics, with practical considerations such as easy access,flexible scheduling, reminder systems, and integration into routine health services are essential in translating acceptance into actual uptake [72]. Indeed, studies show that even parents who express a positive stance toward vaccines may postpone or skip doses due to practical barriers such as physical accessibility and scheduling constraints [73]. Notably, the integration of HPV vaccination into school-based delivery models has proven to be a key determinant of success in many countries, irrespective of their income level [74-75].

## Conclusion

Despite the rapid containment of the COVID19 epidemic, it clearly continues to fuel vaccine hesitancy. Our resultssuggest that introducing new vaccine in a post COVID-era will be connected to a general mistrust (international pharmaceutical industries, experts and health care system), as well as to a need of strong scientific evidence to support safe HPV vaccine uptake concerns about the safety and long-term effects of the vaccines. Addressing the complex issue of mistrust, may employ community-based participatory interventions,as well as culturally sensitive communication strategies,to address concerns and counter misinformation.

## Author Contributions

Conceptualization, Nadia Ben Mansour, Slimane Ben Miled, Judith Mueller, Ahlem Gzara, Emna Hariz and Beya Farah Benzina; methodology, Nadia Ben Mansour, Judith Mueller, Slimane Ben Miled, Emna Hariz, Beya Farah Benzina, Ahlem Gzara, Habiba Ben Romdhane and Saoussen Fray.; software, Nadia Ben Mansour, Beya Farah Benzina and Emna Hariz.; validation, Nadia Ben Mansour, Judith Mueller, Slimane Ben Miled and Habiba Ben Romdhane.; formal analysis,Saoussen Fray, Beya Farah Benzina, and Nadia Ben Mansour.; investigation, Beya Farah Benzina and Saoussen Fray.; resources, Slimane Ben Miled.; data curation, Beya Farah Benzina and Saoussen Fray.; writing—original draft preparation, Nadia Ben Mansour, Saoussen Fray and Beya Farah Benzina.; writing—review and editing, Nadia Ben Mansour, Beya Farah Benzina, Slimane Ben Miled, Habiba Ben Romdhane, Judith Mueller; visualization, Nadia Ben Mansour.; supervision, Nadia Ben Mansour and Slimane BenMiled.; project administration, Slimane Ben Miled and Beya Farah Benzina.; funding acquisition, Slimane Ben Miled . All authors have read and agreed to the published version of the manuscript.”

Special thanks are extended to Professor Mohamed Douagi, General Director of the Office National de la Famille et de la Population, as well as to the coordinators Sameh Ghozzi, LazharNemri, Hatem Lahouimil, Mourad Rjab, Najeh Makni, and Samira Hamdi, for their valuable contributions to the implementation of the study.

## Institutional Review Board Statement

The study was conducted in accordance with the Declaration of Helsinki, and approved by the Institutional Review Board of Pasteur Institute Tunis (2023/14/I/V2, 13 December 2024).

## Informed Consent Statement

Informed consent was obtained from all subjects involved in the study.

## Data Availability Statement

Due to ethical and privacy considerations, the qualitative data (interview transcripts) are not publicly available, as they may contain information that could lead to participant identification. Anonymized excerpt supporting the findings are included in the article. Additional data may be available from the corresponding author upon reasonable request and subject to ethics committee approval.

## Acknowledgments

During the preparation of this manuscript/study, the authors used ChatGPT,free version and Deepl for the purposes of language editing and grammar correction. The authors have reviewed and edited the output and take full responsibility for the content of this publication

